# Evolutionary tree balance predicts disease-free survival in the TRACERx non-small cell lung cancer cohort

**DOI:** 10.1101/2025.11.22.25340797

**Authors:** Kimberley Verity, Robert Noble

## Abstract

Methods for quantifying and classifying modes of tumour evolution promise to enable more personalised prognostic forecasting and treatment optimisation. We recently developed an approach to quantifying evolutionary tree shape that makes full use of the information in tumour clone trees including clone sizes, genetic distances, and phylogenetic relationships. Here, by applying this approach to data from the TRACERx non-small cell lung cancer cohort, we show that clone tree balance predicts disease-free survival after controlling for cancer stage. The association is robust to the omission of rare clones and to the absence of clone sizes or genetic distances. Tree balance outperforms previously proposed evolutionary indices, suggesting that the evenness of evolutionary branching may be more important than the extent of intratumour heterogeneity for determining clinical outcomes.

## 1 Introduction

Prognosis and treatment decisions in non-small cell lung cancer and many other cancer types are primarily based on how far the tumour has spread (stage) and morphological features (grade) [1]. While this system is broadly effective, it lacks precision and fails to account for the dynamic nature of tumour evolution. Proposals to enable more personalised prognostic forecasting include classifying tumours according to their evolutionary and ecological features [2–4].

Although numerous indices have been proposed for summarising the size and shape of evolutionary trees and similar structures [5–7], most studies seeking to develop evolutionary biomarkers in cancer have focused on describing intratumour heterogeneity (ITH) in relatively simple terms. ITH has been associated with tumour progression and therapeutic resistance in multiple cancer types [8]. The TRACERx renal consortium found in their own study and in the larger TCGA kidney cancer cohort that when tumours have low genomic instability, low tumour ITH correlates with longer progression-free and overall survival times [9]. After adjusting for known prognostic variables, including stage and grade, both ITH and genomic instability remained significant predictors in the TCGA kidney cancer cohort but not in their own cohort [9]. In the larger TRACERx non-small cell lung cancer (NSCLC) cohort that we reanalyse here, disease-free survival has been shown to be related to the ITH of somatic copy number alterations but not to mutational ITH [10]. In a multivariate Cox model controlling for stage and other clinical variables, the ITH of somatic copy number alterations was no longer predictive [10]. Measures of heterogeneity have also been found to be predictors of clinical outcome in prostate cancer and in a pan-cancer analysis across 28 cancer types [11, 12].

A complementary but comparatively under-investigated summary statistic is tree balance: the degree to which internal nodes split their descendants into equally sized subtrees [13]. Balance indices capture a fundamental aspect of tree shape that characteristically varies between evolutionary processes. Tree balance can thus be used to infer parameter values or to compare empirical trees with those generated by mathematical models [13–17].

Among the many tree balance indices that have been developed [18], the most popular include Sackin’s index, which sums the distances between the terminal nodes (or leaves) and the root [19], and Colless’ index, which sums the absolute differences between the number of leaves descending from the right and left branches of each internal node [20]. Such conventional tree balance indices were designed to be applied to species trees and are less well suited to comparing the shapes of tumour phylogenies. Weaknesses include not accounting for node sizes (corresponding to clone abundances) and branch lengths (genetic distances or divergence times); sensitivity to the inclusion or omission of rare types; and inapplicability to non-bifurcating trees [21, 22].

We recently defined a new system of indices that capture tree balance, diversity, and other aspects of tree shape while avoiding the limitations of prior approaches [22]. Our indices account for both node sizes and branch lengths and are robust to small changes in either attribute. They assign interpretable values to all trees and enable a meaningful comparison of the shapes of any pair of trees. Consider the two tumour trees shown in Figure 1. Mutational ITH – previously used in the TRACERx NSCLC study – assigns the same value 0.03 to both trees and so cannot distinguish between them. Our tree balance and diversity indices, ^1^*J*_*N*_ and ^1^*D*_*L*_, capture the obvious differences in shape (Figure 1a: ^1^*J*_*N*_ = 0.77 and ^1^*D*_*L*_ = 1.70; Figure 1b: ^1^*J*_*N*_ = 1 and ^1^*D*_*L*_ = 1).

**Figure 1:**
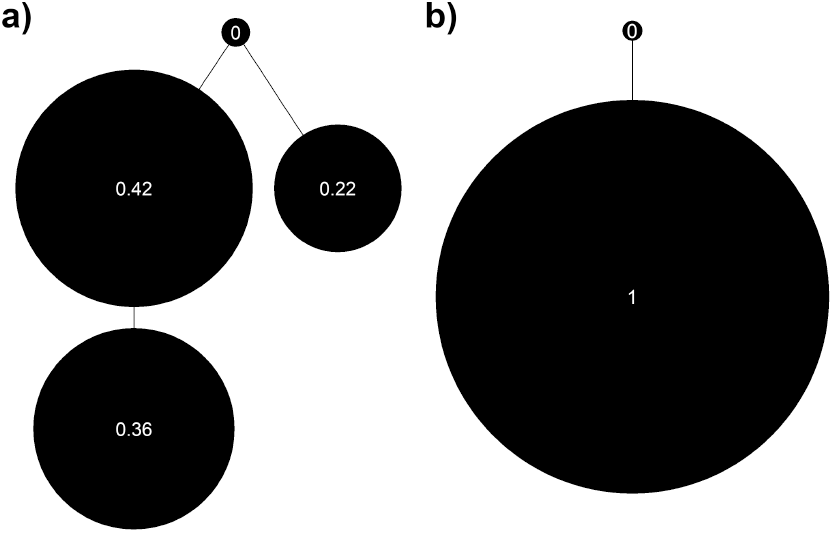
Tumour trees illustrating a case where mutational ITH is identical and cannot distinguish between the trees but ^1^*J*_*N*_ can. a) tumour ID CRUK0254, mutational ITH = 0.03 and ^1^*J*_*N*_ = 0.77, b) tumour ID CRUK0092, mutational ITH = 0.03 and ^1^*J*_*N*_ = 1. Branch lengths are arbitrary and node sizes are proportional abundances.

Here we show that our new tree balance index outperforms measures of intratumour heterogeneity in predicting disease-free survival in non-small cell lung cancer.

## 2 Results

### 2.1 New tree shape indices predict disease-free survival

We apply tree shape indices to 392 phylogenetic trees reconstructed for tumours from patients from the TRACERx non-small cell lung cancer cohort. Trees range from having 1 to 10 leaves, and the average number of total nodes in a tree is 10.34. Node sizes correspond to the overall proportion of tumour cells belonging to each clone, and branch lengths correspond to genetic distance. Figure 2 shows four examples.

**Figure 2:**
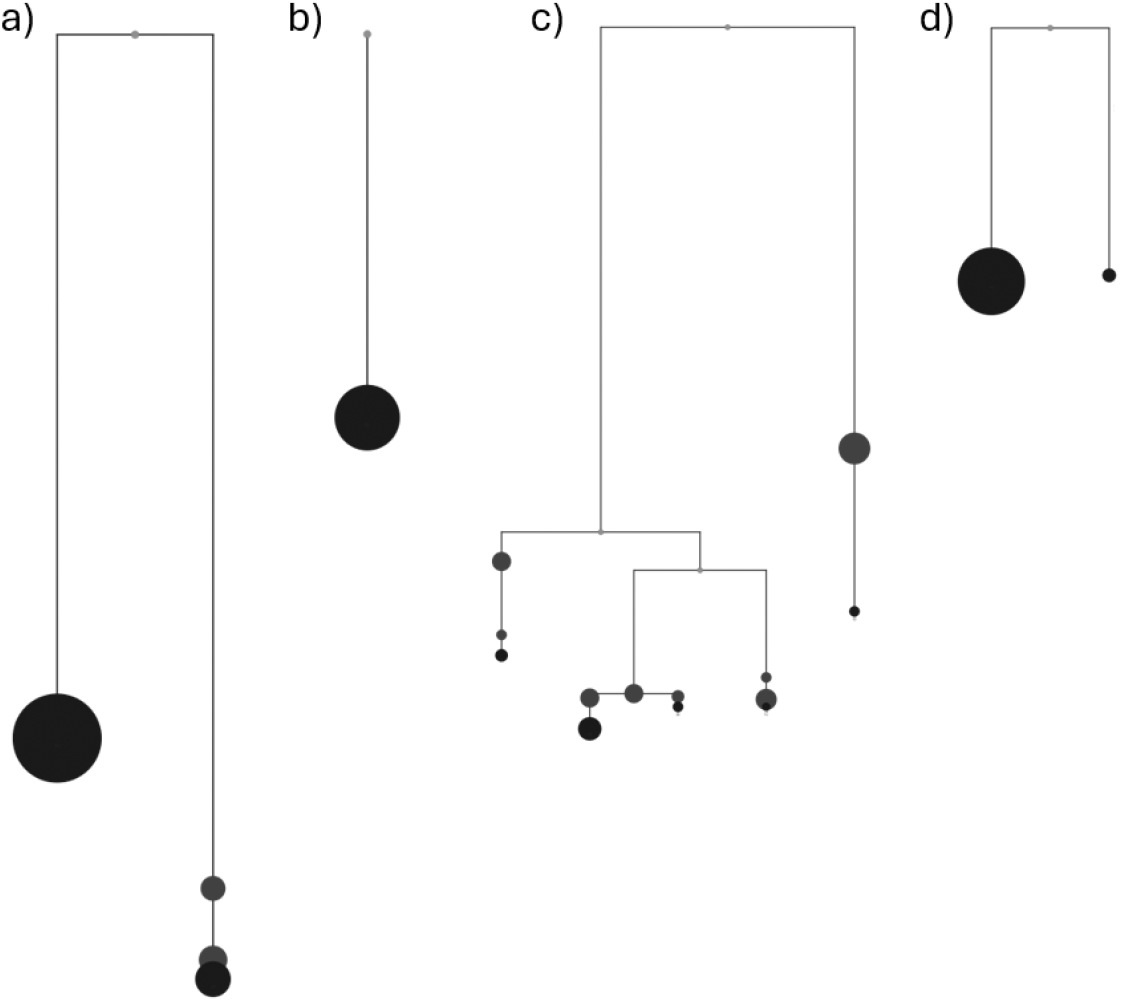
a-b) Completely balanced (^1^*J*_*N*_ = 1) and c-d) unbalanced (^1^*J*_*N*_ < 0.73) trees shown with proportional abundances (not consistent between plots) and branch lengths (consistent between plots). Red nodes either have zero abundance or are the root node. Tumour IDs a) CRUK0027, b) CRUK0061, c) CRUK0284 and d) CRUK0756. Trees were drawn using PhyloWeaver [23].

We initially investigate the relationship between disease-free survival (DFS) and three of our new indices: ^1^*D*_*N*_,^1^ *D*_*L*_ and ^1^*J*_*N*_. Treating the indices as continuous variables, we find a significant association between DFS and all three indices (^1^*D*_*N*_ hazard ratio (HR) = 1.21, 95% confidence interval (CI) = 1.06-1.39, ^1^*D*_*L*_ HR = 1.21, 95% CI = 1.05-1.38, and ^1^*J*_*N*_ HR = 0.79, 95% CI = 0.69-0.90. When stating hazard ratios for continuous variables, the variables have been scaled such that one unit change is equivalent to one standard deviation, this allows a fair comparison of the effect of variables across different scales.) In a multivariable Cox proportional hazards model accounting for all three indices, only ^1^*J*_*N*_ predicts DFS (HR = 0.79, 95% CI = 0.69-0.91). We also looked at the non-normalised version of ^1^*D*_*L*_ but found no significant relationship with DFS (HR = 1.09 95% CI = 0.95-1.26).

Using the indices to assign patients to three categories (see Methods), we find a significant association between DFS and ^1^*J*_*N*_ in all pairwise comparisons (Figure 3c). High ^1^*D*_*N*_ and ^1^*D*_*L*_ values also predict shorter DFS when compared to low or intermediate values (Figure 3a,b). As the tree balance index ^1^*J*_*N*_ performed best in these analyses we will henceforth focus on this one index.

**Figure 3:**
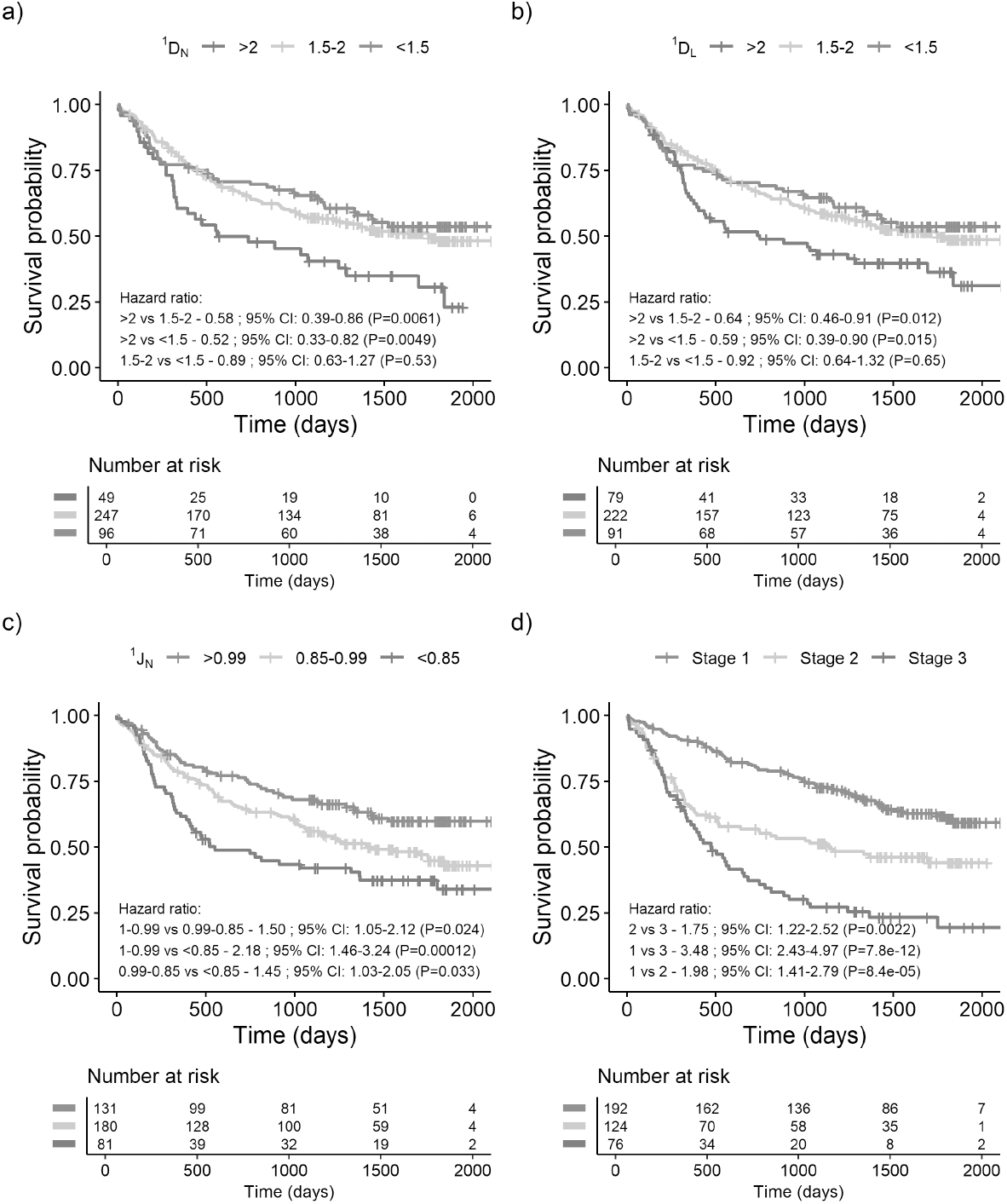
Survival curves showing the difference in disease-free survival for tumours based on tree shape indices a) ^1^*D*_*L*_, b) ^1^*D*_*N*_, c) ^1^*J*_*N*_ and d) the stage.

Of the 392 trees 23 have ^1^*J*_*N*_ = ^1^*D*_*L*_ = ^1^*D*_*N*_ = 1, 18 are linear trees and 5 contain only the root node and so we defined the indices to be 1. Removing these trees has minimal effect on the results.

### 2.2 Associations between tree balance and clinical features

^1^*J*_*N*_ is significantly lower in cancer stages 2 and 3 than in stage 1 (1 vs 2: ^1^*J*_*N*_ difference = 0.041, *P* = 0.001; 1 vs 3: 0.049, *P* = 0.001; Supplementary Figure 9) but does not differ between stages 2 and 3 (^1^*J*_*N*_ difference = 0.008, *P* = 0.86). Tree balance also correlates with other disease and treatment factors including whether the patient had lymphovascular invasion, the surgery type, and whether the patient had adjuvant treatment. Such clinical factors are linked through clinical decision-making pathways because treatment and surgical decisions are largely determined by tumour stage. We found no significant associations between tree balance and patient attributes such as age and smoking history, nor between tree balance and any particular somatic mutation.

### 2.3 Tree balance remains prognostic after controlling for stage

As stage is also predictive of DFS (Figure 3d) and is only weakly associated with ^1^*J*_*N*_, we next investigated whether stage and ^1^*J*_*N*_ perform better in combination than either does alone. When we split the cohort according to stage, we observe a significant relationship between tree balance and DFS within stage 2 when comparing the most and the least balanced trees (Figure 5b, HR = 2.12, 95% CI = 1.07-4.21). After controlling for stage in multivariate Cox models, tree balance remains a significant predictor of DFS when comparing the most and least balanced trees (HR = 1.6, 95% CI = 1.07-2.4; Figure 4). The hazard ratios for tree balance change little when we control for grade as well as stage but are no longer significantly different from unity (Figure 10). The latter analysis suffers from much reduced sample size (191 patients and 86 events) because grade information is available only for lung adenocarcinomas.

**Figure 4:**
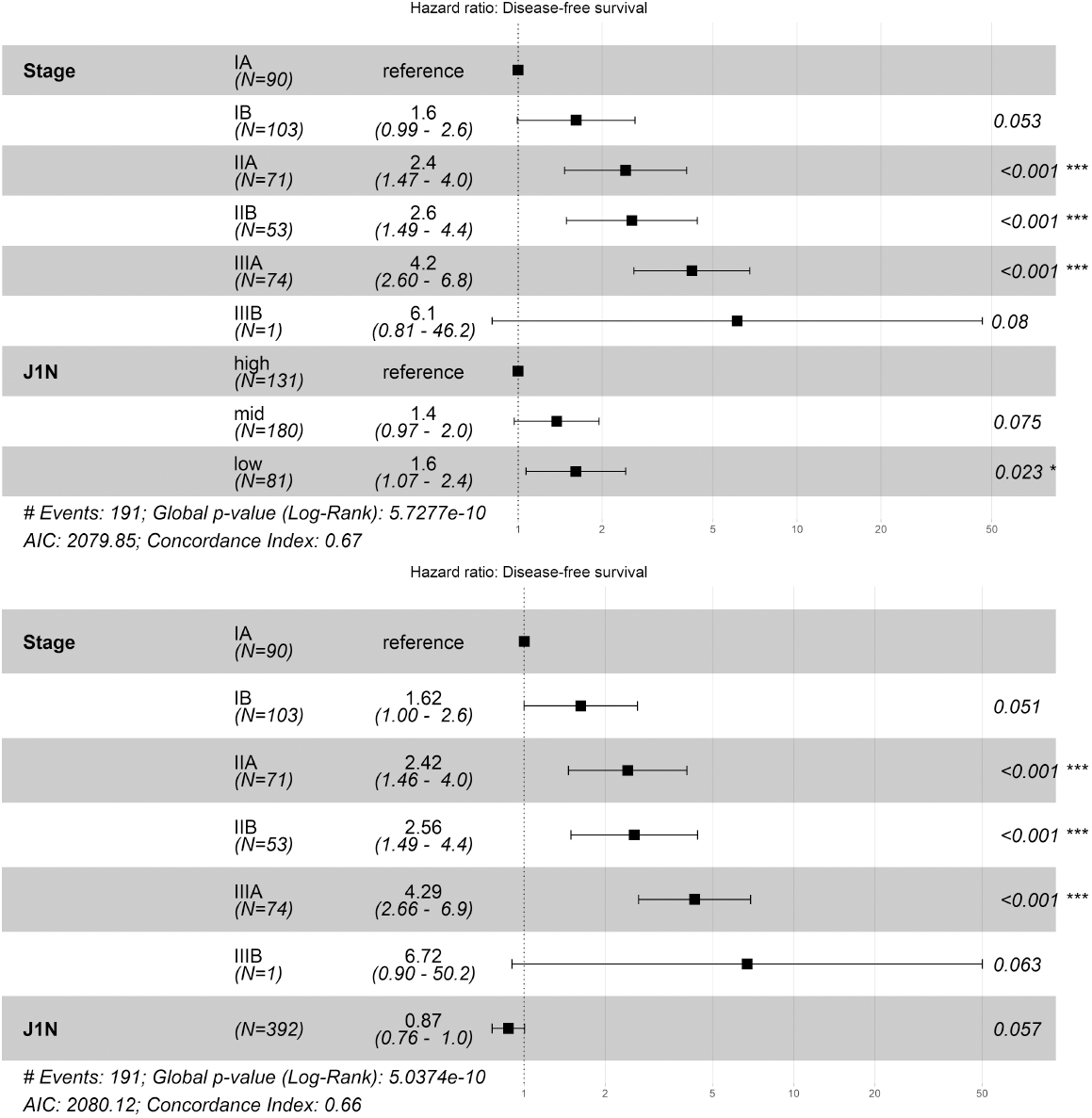
Multi-variable Cox proportional hazard models containing stage and tree balance, ^1^*J*_*N*_, a) split into intervals, and b) as a continuous variable. The HR 95% CIs are shown in brackets and by the error bars. The asterisks indicate the *P* value ranges, where \**P <* 0.05, \*\**P* < 0.01, \*\*\**P* < 0.001.

**Figure 5:**
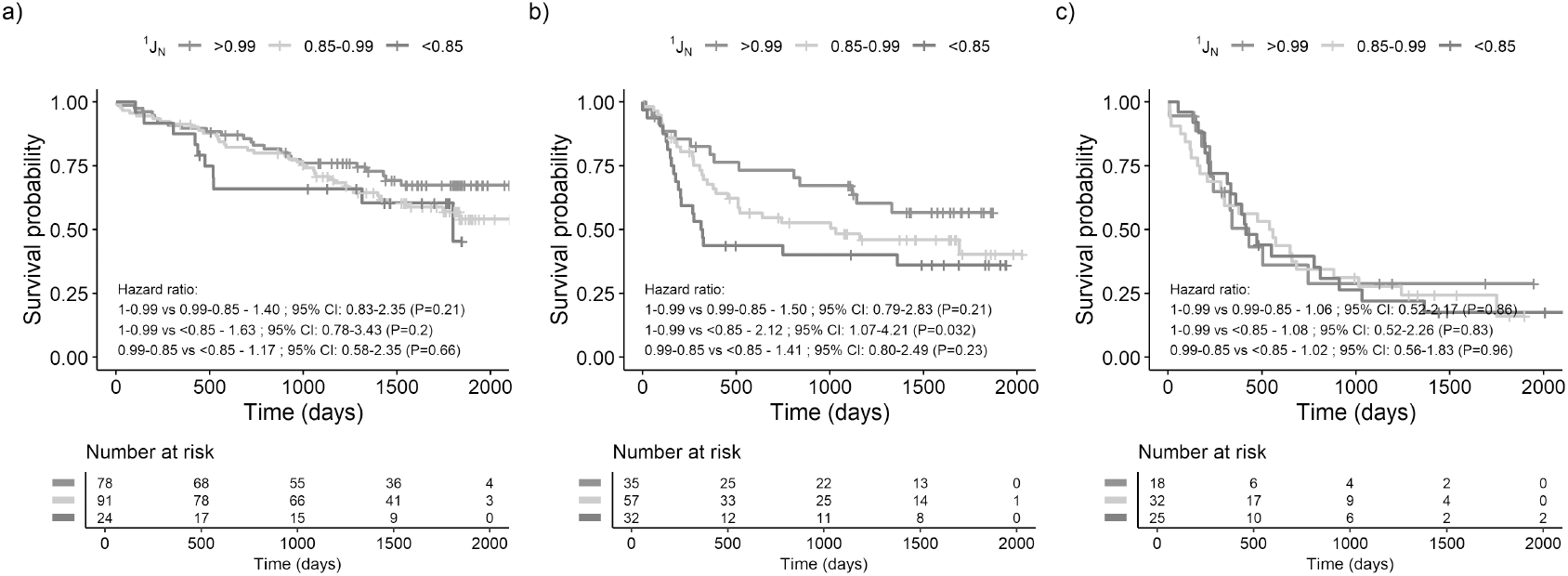
Survival curves showing the difference in DFS for tumours based on their phylogenetic tree balance, ^1^*J*_*N*_, when split by stage where, a) stage 1, b) stage 2 and c) stage 3.

### 2.4 Results are insensitive to the omission of rare clones

To test whether tree balance remains predictive when applied to poorer quality data, we examined the effect of omitting rare clones from the TRACERx trees. We identified nodes corresponding to tumour clones with low proportional abundances and then merged each such node with its parent (Figure 6; Figure 11). Merging nodes with abundances (including abundances of any descendant clones) less than 1%, 5% and 10% of the total abundance in the tree reduced the mean number of nodes per tree by 0.09 %, 8.8% and 25.7 %, respectively. For all three tolerance values, the results varied little. DFS remained significantly related to ^1^*J*_*N*_ treated either as a continuous variable (HR = 0.79, 95% CI = 0.69-0.90; HR = 0.81, 95% CI = 0.71-0.92; HR = 0.84, 95% CI = 0.74-0.96; for thresholds 1% and 5% and 10% respectively) or (in all but one of the pairwise comparisons per threshold) as a categorical variable (Figure 7). The removal of rare clones typically increases tree balance, resulting in more patients being initially assigned to the high-^1^*J*_*N*_ category (Figure 7). As we increasingly remove rare types, the difference in DFS between the high- and medium-^1^*J*_*N*_ categories diminishes, while the difference between the medium- and low-^1^*J*_*N*_ categories increases. Similar results pertain for multivariate models controlling for stage (HR = 0.87 95% CI = 0.76-1.00; HR = 0.89 95% CI = 0.78-1.02; HR = 0.92 95% CI = 0.80-1.05 for thresholds 5% and 10% respectively, with ^1^*J*_*N*_ as a continuous variable).

**Figure 6:**
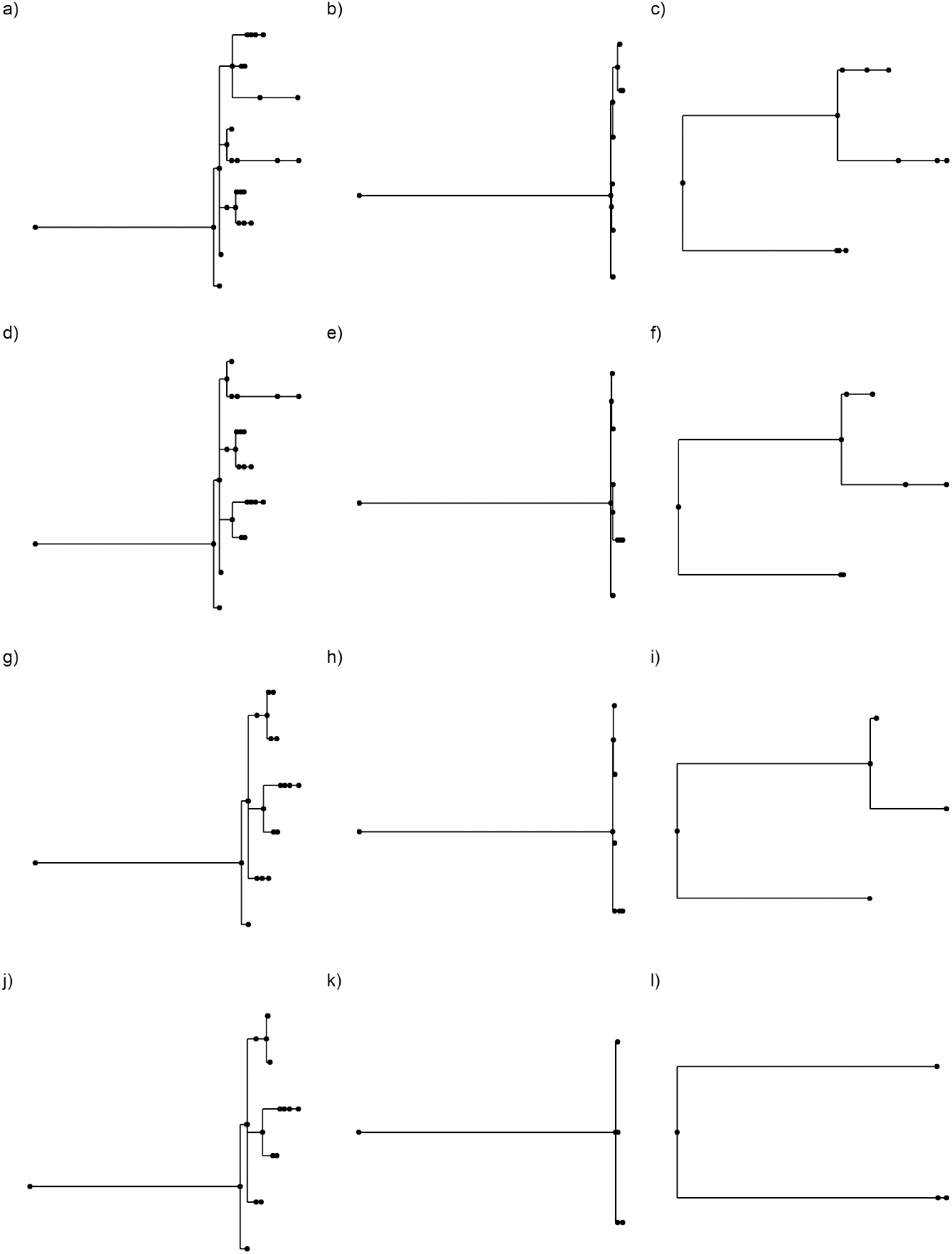
Three tumour trees at different levels of coarse-graining. a-c) the original trees, d-f) 1%, g-i) 5% and j-l) %10. (Tumour IDs CRUK0065, CRUK0462 and CRUK0496 respectively). Trees are shown with branch length only.

**Figure 7:**
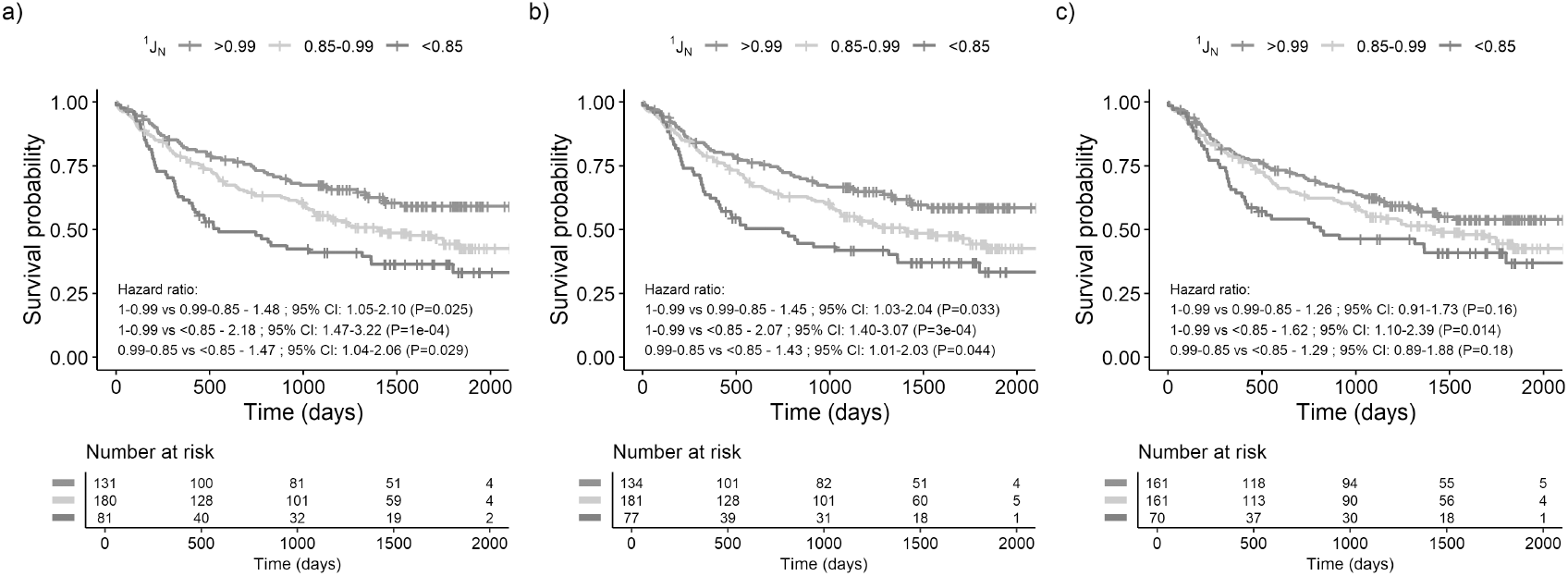
Survival curves showing the difference in DFS for tumours for varying amounts of coarse-graining, a) 1%, b) 5% and c) 10%.

### 2.5 Results are robust to absence of clone size and branch length data

We next compared the predictive power of ^1^*J*_*N*_ to that of three variants of our tree balance index that account for branch lengths but not node sizes (^1^*J*_*N,a*_); account for node size but not branch lengths (^1^*J*_*N,b*_); or account for neither branch lengths nor node sizes (^1^*J*_*N,c*_). In the first and last cases (consistent with the convention for cladograms [21]) we assigned size one to the leaves of each tree and size zero to the internal nodes. For fairer comparison with ^1^*J*_*N*_, we adjusted the lower cutpoint boundary for each alternative balance index to maintain approximately 80 trees in the low-balance category, while keeping the upper cutpoint unchanged (see Figure 12 for results using the ^1^*J*_*N*_ lower cutpoint for all indices). As categorical variables, all variants of our tree balance index give similar results (Figures 8a-c). As continuous variables, both individually and in multivariate models with stage, the variants perform similarly but ^1^*J*_*N,b*_ performs the best (^1^*J*_*N,a*_: HR = 0.78, HR = 0.87, ^1^*J*_*N,b*_ HR = 0.76, HR = 0.84, ^1^*J*_*N,c*_ HR = 0.77, HR = 0.86 without and with stage respectively). For ^1^*J*_*N,b*_, 75% of trees remain in the same category as for ^1^*J*_*N*_; for ^1^*J*_*N,a*_, 70% remain in the same category; and for ^1^*J*_*N,c*_ the consistency is 64%. These results suggest that it is more important to account for node sizes (tumour clone sizes) than for branch lengths (genetic distances between clones).

**Figure 8:**
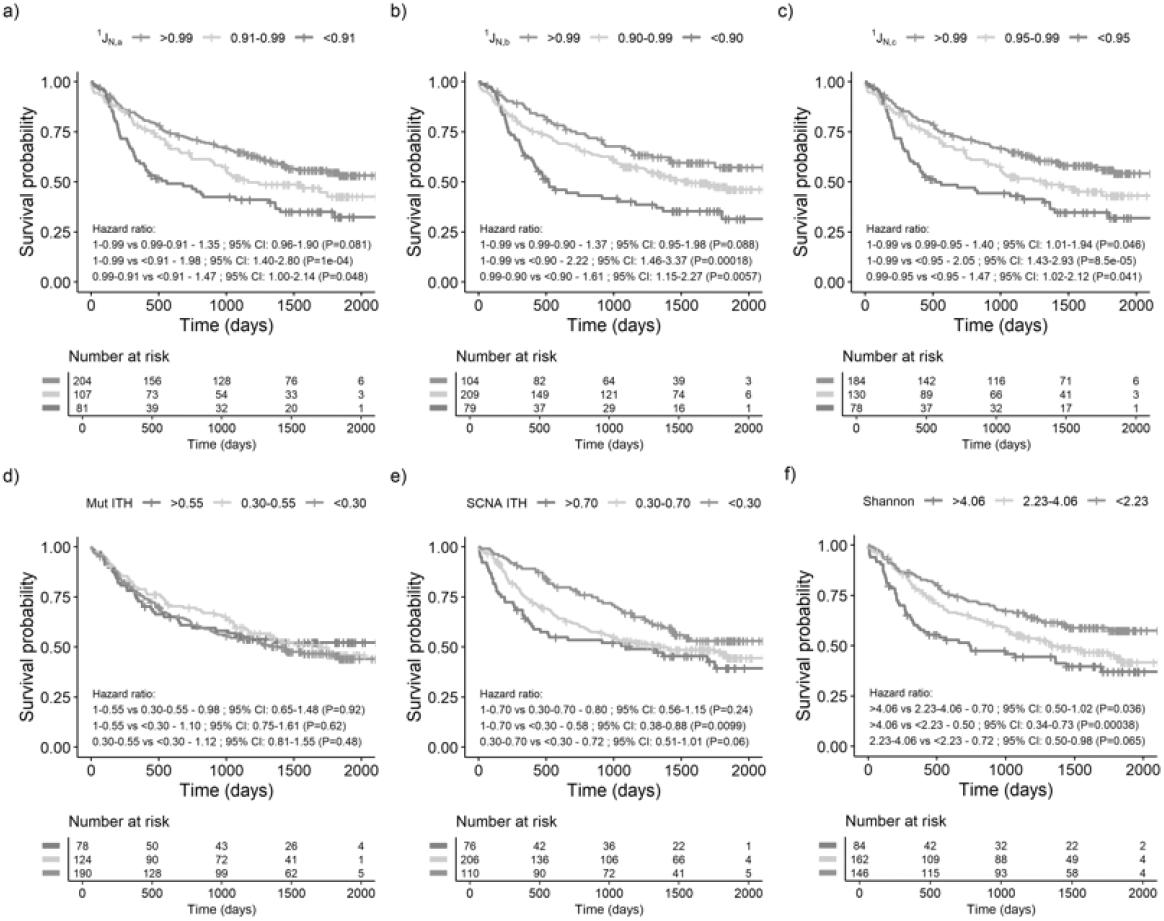
Survival curves showing the difference in disease-free survival for tumours based on alternative indices. a-c) are alternative versions of our tree balance index, where a) ^1^*J*_*N,a*_ accounts for branch lengths but leaves are assumed to have equal abundance and internal nodes have size zero, b) ^1^*J*_*N,b*_ accounts for node sizes but not branch lengths, and c) ^1^*J*_*N,c*_ accounts for neither node sizes or branch lengths. d) Mutational ITH is the percentage of mutations that are subclonal. e) Somatic copy number alteration (SCNA) ITH is the fraction of aberrant genome with subclonal SCNAs, both d and e are taken from [10]. f) Shannon diversity in units of effective types calculated on leaves only.

The variant indices as continuous variables appear to perform slightly better than ^1^*J*_*N*_, with smaller hazard ratios that are more significant. However, this is not true when the indices are transformed to categorical variables. Overall, no variants outperform ^1^*J*_*N*_. Without stage, only ^1^*J*_*N,b*_ outperforms ^1^*J*_*N*_ in one comparison having a larger HR that is more significant, the others do not outperform ^1^*J*_*N*_ in both cases - larger HR that is also more significant - in any comparison (Figures 3c and 8). With stage, ^1^*J*_*N,b*_ performs very similarly to ^1^*J*_*N*_, and ^1^*J*_*N*_ outperforms the other variants (Figures 4 and 13).

Coarse-graining with respect to either abundances or branch lengths (at 10% and 1.5% level respectively, which removes approximately the same number of branches) leads to ^1^*J*_*N,a*_ no longer being significant (HR = 0.90 95% CI = 0.78-1.02 and HR = 0.94 95% CI = 0.82-1.08 respectively). Coarse-graining with respect to branch lengths at the 1.5% level leads to ^1^*J*_*N,c*_ no longer being significant (HR = 0.93 95% CI = 0.69-1.06). For both types of coarse graining, ^1^*J*_*N*_ and ^1^*J*_*N,b*_ have HRs that change little and remain significant. However, as categorical variables, ^1^*J*_*N,b*_ performs worse and loses significance first.

### 2.6 New indices outperform prior evolutionary indices

Next, we compare the predictive power of our tree shape indices to three alternative measures of intratumour heterogeneity. We investigate mutational ITH and SCNA ITH, which were used in previous analyses of the TRACERx non-small cell lung cancer cohort [10, 24], and the Shannon diversity. Mutational ITH is the percentage of subclonal mutations, and somatic copy number alteration (SCNA) ITH is the fraction of aberrant genome with SCNAs.

For mutational ITH, we find no significant relationship with DFS when the ITH index is treated as either a categorical variable (Figure 8d) or as a continuous variable. For ITH in terms of somatic copy number alterations (SCNA) as a continuous variable, we find a significant relationship with DFS (HR = 1.19 95% CI = 1.03-1.37). Treating SCNA ITH as a categorical variable, we find a significant difference in DFS when comparing the high- and low-value categories (Figure 8e). But in a multivariable model controlling for stage, SCNA ITH is no longer significantly associated with DFS, consistent with previous results [10].

For the Shannon diversity calculated on the leaves of the trees, as a continuous variable, we find a significant relationship with DFS (HR = 1.28 95% CI = 1.12-1.46). As a categorical variable, we find a significant relationship with DFS in comparisons with the high-value categories (Figure 8f). However, in multivariable models controlling for stage, the Shannon diversity is no longer significantly associated with DFS. We also calculate the Shannon diversity on all nodes in the tree, finding that this is never significantly associated with DFS. This analysis confirms that our tree balance index ^1^*J*_*N*_ outperforms prior evolutionary indices for predicting DFS in this cohort.

## 3 Discussion

We have shown that in the TRACERx non-small cell lung cancer cohort, there is a significant relationship between disease-free survival (DFS) and aspects of tumour clone tree shape, including the effective out-degree (^1^*D*_*N*_), the effective number of maximally distinct leaves (^1^*D*_*L*_), and tree balance (^1^*J*_*N*_). Among these three indices, tree balance performs the best. Multivariable Cox models with both stage and tree balance showed that tree balance remains a significant predictor after accounting for cancer stage. The main advantage of tree balance is in stratifying patients in stage 2.

We demonstrated that the removal of rare nodes at small tolerance values has very little effect on our results. As the number of nodes removed increases, our balance index begins to lose power, however, for all three levels of coarse-graining, the relationship between tree balance and DFS remained significant. Therefore we have shown the robustness of our method to the omission of rare types.

We find that tree topology accounts for the main diagnostic signal, with branch length and abundance data being unnecessary when fine-scale structure is intact, as is the case here with the high-quality data used to generate the trees. Branch lengths become important when we consider the categorical stratification of the coarse-grained data, suggesting that branch length information enhances discrimination between clinically distinct groups once short branches are removed. These results imply that, in well-resolved data, topology alone captures most of the prognostic information, but ^1^*J*_*N*_, which accounts for abundances and branch lengths - clone size and genetic distance here - provides robustness and improved patient stratification as data quality decreases.

Evolutionary and ecological processes in cancer are known to be important, yet there is a need to develop methods that map the differences in tumour evolution into information that matters for patient outcomes [4]. Tree balance has long been used to study evolutionary processes [13], primarily in systematic biology but increasingly in other research areas such as cancer. It can detect branching rate heterogeneity in lineage-tracing data [16], and is associated with immunotherapy response in colorectal cancer [17]. These results, along with our finding that tree balance is significantly associated with DFS, suggest that the tree balance captures clinically relevant aspects of tumour evolution. Ultimately, highlighting tree balance as an emerging and informative lens for studying cancer evolution.

To understand why we find tree balance to have such prognostic power, we consider the evolutionary dynamics that may give rise to this pattern. Clonal diversity has been shown to predict tumour growth and outcome primarily as a proxy for intrinsic biological factors such as mutation rate and clonal turnover [25], while spatial models indicate that tumour architecture and cell dispersal dynamics influence these same processes [26]. Tree balance may therefore act as a proxy for the combined influence of biological and ecological constraints on tumour evolution. The better prognostic performance of tree balance compared with diversity-based indices may suggest that the evenness of evolutionary branching, rather than the effective out-degree or number of leaves, is more important for determining clinical outcomes.

In conclusion, we have used a general set of indices quantifying aspects of tree shape and shown that they have a significant relationship with DFS. Moreover, combining tree balance and stage leads to better patient stratification than stage alone. This demonstrates that tree balance captures clinically relevant aspects of tumour evolution. The mechanisms underlying this association remain unclear. Investigating these mechanisms will be essential to determine whether tree balance can be used as a prognostic tool.

## 4 Methods

### 4.1 Tree shape indices

Our indices for quantifying tree shape have been previously described [22]. Briefly, ^1^*D*_*N*_ quantifies the average effective out-degree or, more informally, the “bushiness” of the tree; ^1^*D*_*L*_ is a diversity index that accounts for phylogenetic relatedness; and ^1^*J*_*N*_ is a tree balance index. All three indices account for tree topology, node sizes (here corresponding to subclone population sizes) and branch lengths (genetic distance). The two *D* indices can take any positive value, whereas ^1^*J*_*N*_ varies between 0 (minimally balanced) and 1 (perfectly balanced).

### 4.2 TRACERx data

The TRACERx 421 cohort contains 421 patients recruited across 19 hospital sites in the United Kingdom. The recruitment was broadly representative of an early-stage operable non-small cell lung cancer (NSCLC) population in the UK according to ethnicity, age, sex and smoking status [10]. The 421 patients had 432 genomically independent tumours: 248 lung adenocarcinomas (LUAD); 138 lung squamous cell carcinomas (LUSCs); and 46 ‘other’ NSCLC subtypes. Pathological staging was available for all tumours but tumour grading was only available for LUADs [10, 27]. Tumour phylogenetic trees were reconstructed from multiregion whole-exome sequencing (WES) data using the CONIPHER computational framework to infer the evolutionary relationships between tumour clones [10, 28]. Nodes in the trees correspond to genetically defined clones, comprising tumour cells that are identical by descent in their somatic mutation history. The branching structure captures ancestral relationships between clones, where descendant clones inherit the mutational profile of their parent clones but may also acquire additional alterations or lose ancestral mutations through copy-number changes or other genomic events.

Phylogenetic trees could be reconstructed for 401 tumours; 9 were excluded from the analysis. One patient had two synchronous primary tumours, one of which was not sequenced. Additionally, for the other patients with synchronous primary tumours we used the tumour with the highest stage; this removed a further 8 trees. For branch lengths, we used the absolute difference in the number of mutations between parent and child clones. Due to the tree construction method that allows for somatic copy number alterations (SCNAs) to remove mutations, child clones could have fewer mutations than their parent, hence the need to use the absolute number. Clone sizes were obtained from CONIPHER using the cancer cell fractions (CCF). 5 of the remaining 392 trees only contained the root node, a case in which our indices are not defined. To include these tumour trees in the analysis we assigned a tree with only the root node to have index values of one. Figure 2 shows four of the tumour trees, a and b are completely balanced and c and d are unbalanced.

### 4.3 Cutpoints for categorical analysis

Choosing cutpoints to group continuous variables is not a simple task and there is not a universally agreed way to do it, and crucially the results of analyses can change drastically if different cutpoints are used [29]. Commonly used cutpoints are splitting around the median and the method of “optimising” the P-value which is equivalent to minimising the P-value. However, just because they are commonly used does not mean they are without their pitfalls. Splitting around the median value gives even group sizes, but other than that, it is as arbitrary as splitting around any other value. The minimum P-value, although it may seem mathematically desirable, has issues from limiting the ability to compare studies, to an inflated type I error rate [29]. Altman et al. demonstrated the associated issues with the minimum P-value method using the example of S-phase fraction as a prognostic marker in breast cancer in [29]. Here we took their recommendation where ‘the choice of cutpoints should be guided by biological reasoning, knowledge of measurement techniques, and simplicity’ [29]. As our indices are mathematical and not biological, we used simplicity when selecting our cutpoints, where we chose the cutpoints such that the groupings made sense based on the index values and also kept reasonable group sizes. Given this, the groupings here may not be optimal, and we include the analysis for each index as a continuous variable to demonstrate the results are not just due to the chosen cutpoints.

## Data Availability

All data produced in the present work are contained in the manuscript

## Supplementary figures

**Figure 9:**
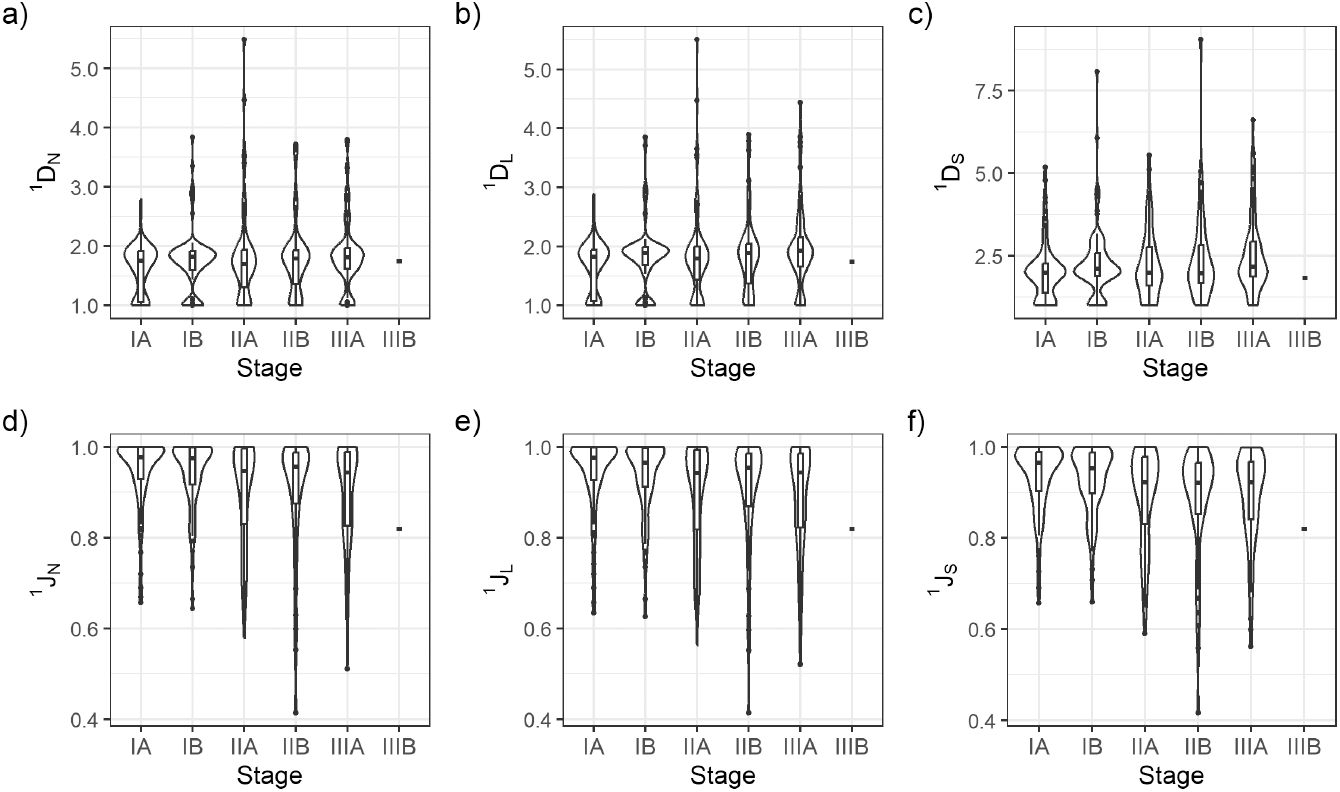
Violin and box plots for indices ^1^*D*_*N*_, ^1^*J*_*N*_, ^1^*D*_*L*_, ^1^*J*_*L*_, ^1^*D*_*S*_, ^1^*J*_*S*_ split based on stage. Stage IIIB contains only two patients and hence is not plotted.

**Figure 10:**
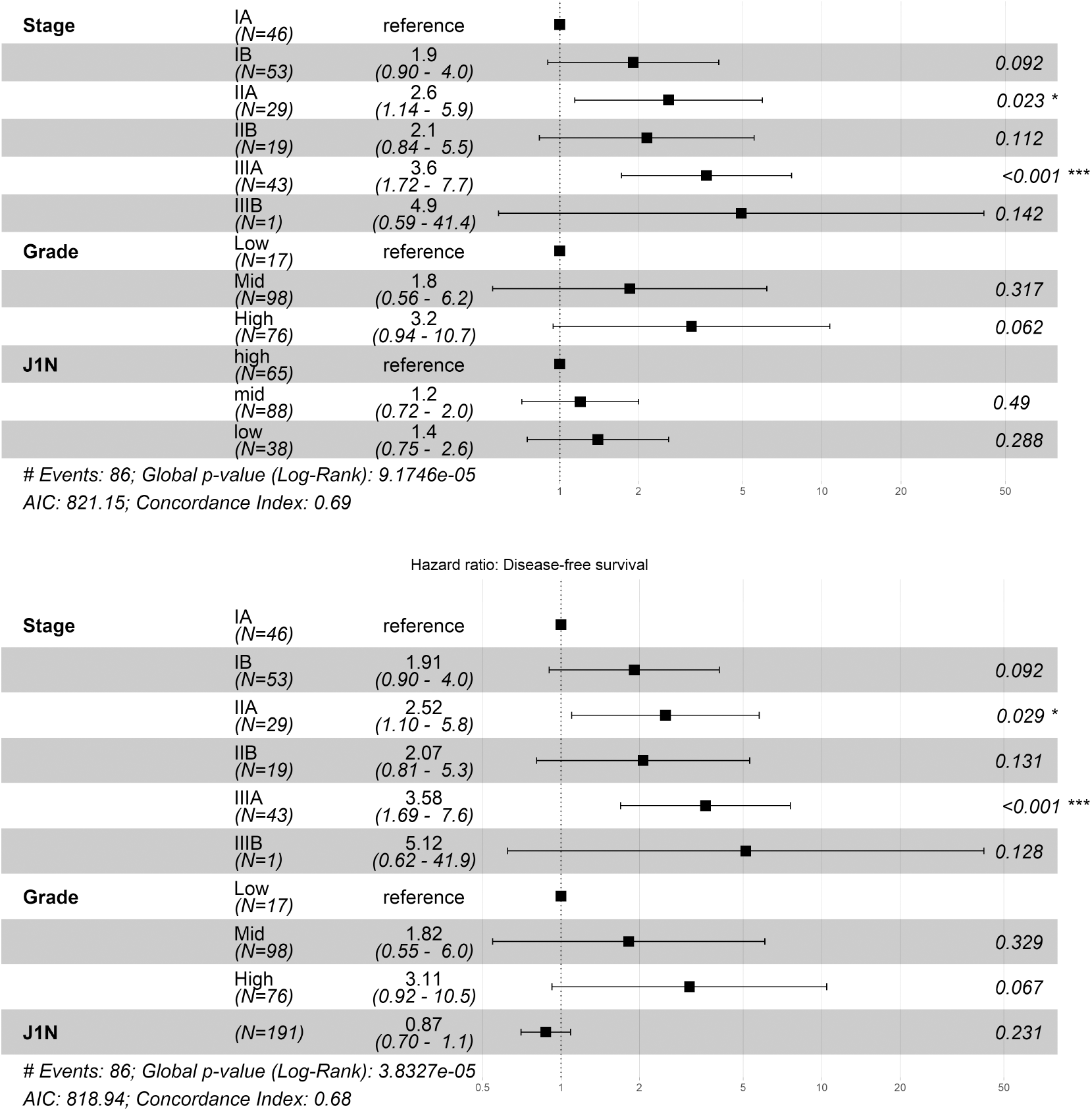
Multi-variable Cox proportional hazard models containing stage, grade and tree balance, ^1^*J*_*N*_, a) split into intervals, and b) as a continuous variable. The HR 95% CIs are shown in brackets and by the error bars. The asterisks indicate the *P* value ranges, where \**P* < 0.05, \*\**P* < 0.01, \*\*\**P* < 0.001.

**Figure 11:**
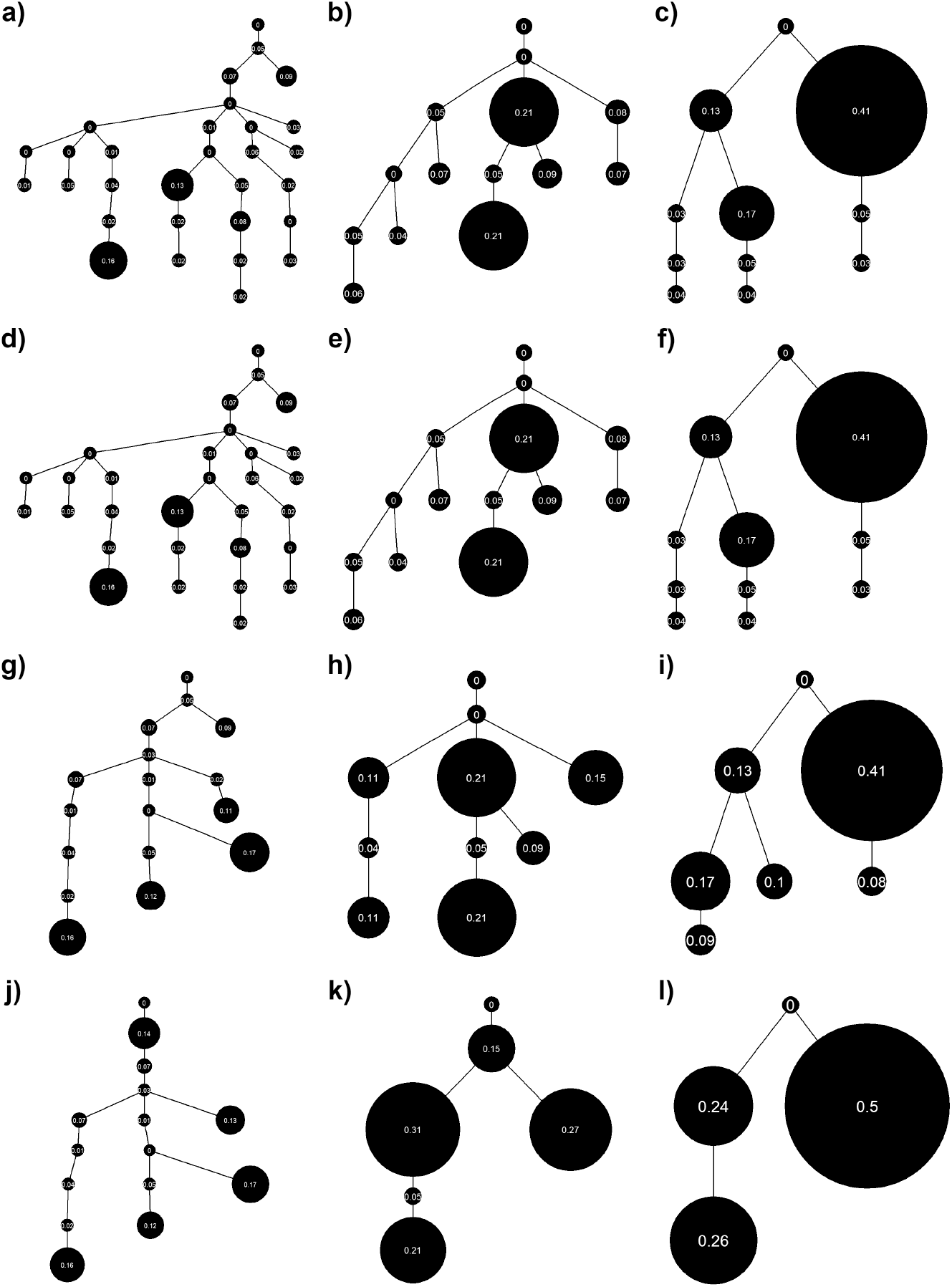
Three tumour trees at different levels of coarse-graining. a-c) the original trees, d-f) 1%, g-i) 5% and j-l) 10%. (Tumour IDs CRUK0065, CRUK0462 and CRUK0496 respectively). Trees are shown with proportional node sizes only (branch lengths are arbitrary).

**Figure 12:**
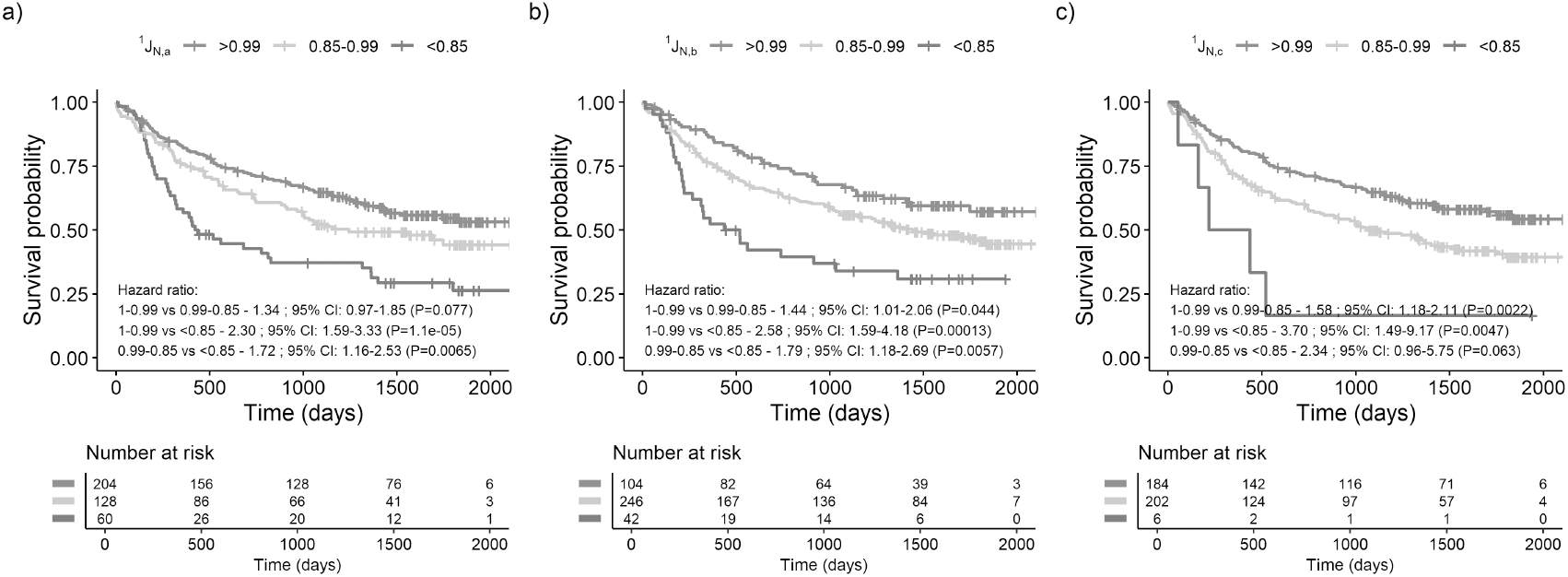
Survival curves showing the difference in DFS for tumours based on alternative tree shape indices with the original cut-points.

**Figure 13:**
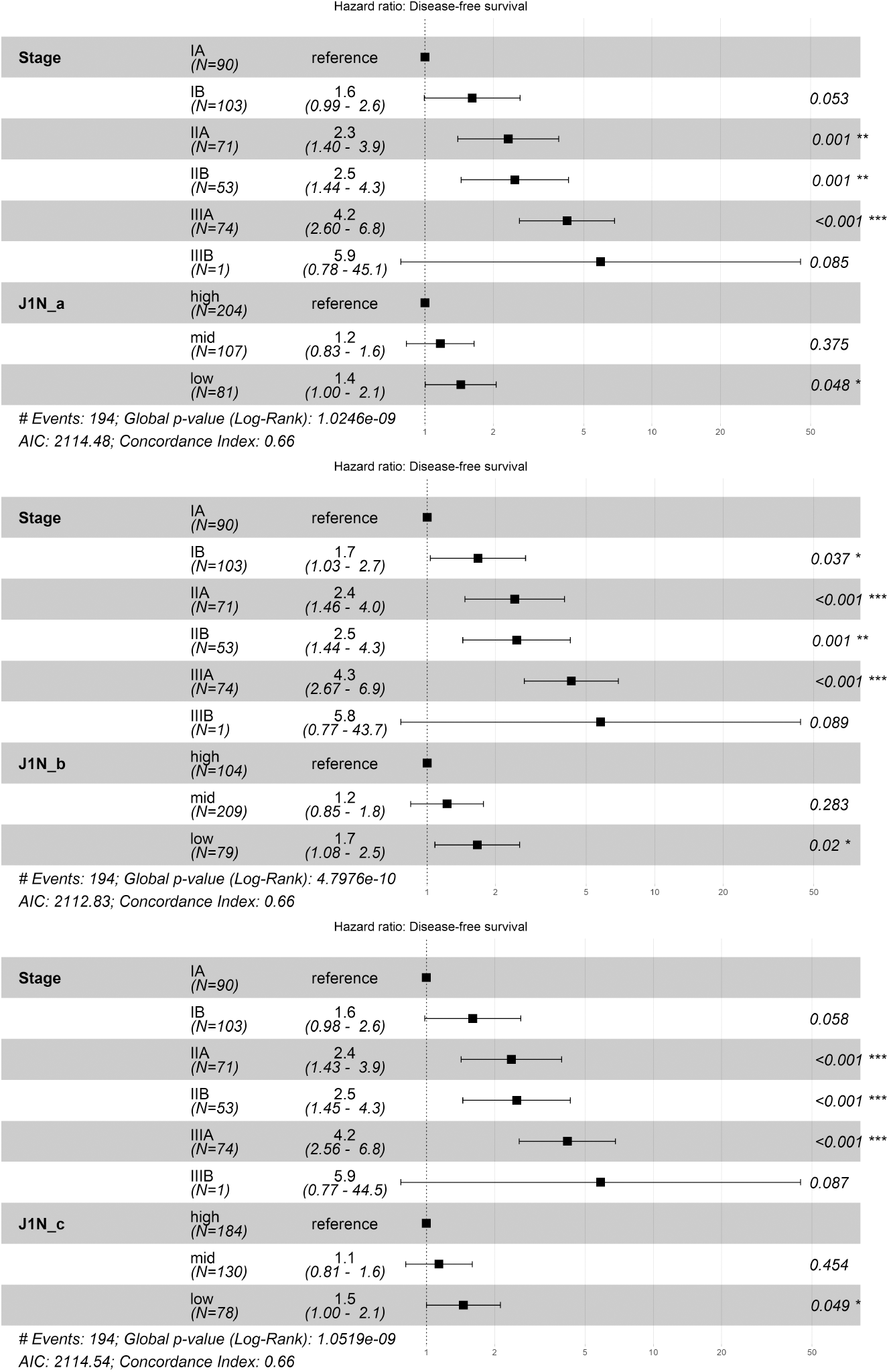
Multi-variable Cox proportional hazard models containing stage and alternative tree balance indices. The adjusted cutpoints used in Figure 8 are the cutpoints used here. The HR 95% CIs are shown in brackets and by the error bars. The asterisks indicate the *P* value ranges, where \**P*< 0.05, \*\**P* < 0.01, \*\*\**P* < 0.001.

**Figure 14:**
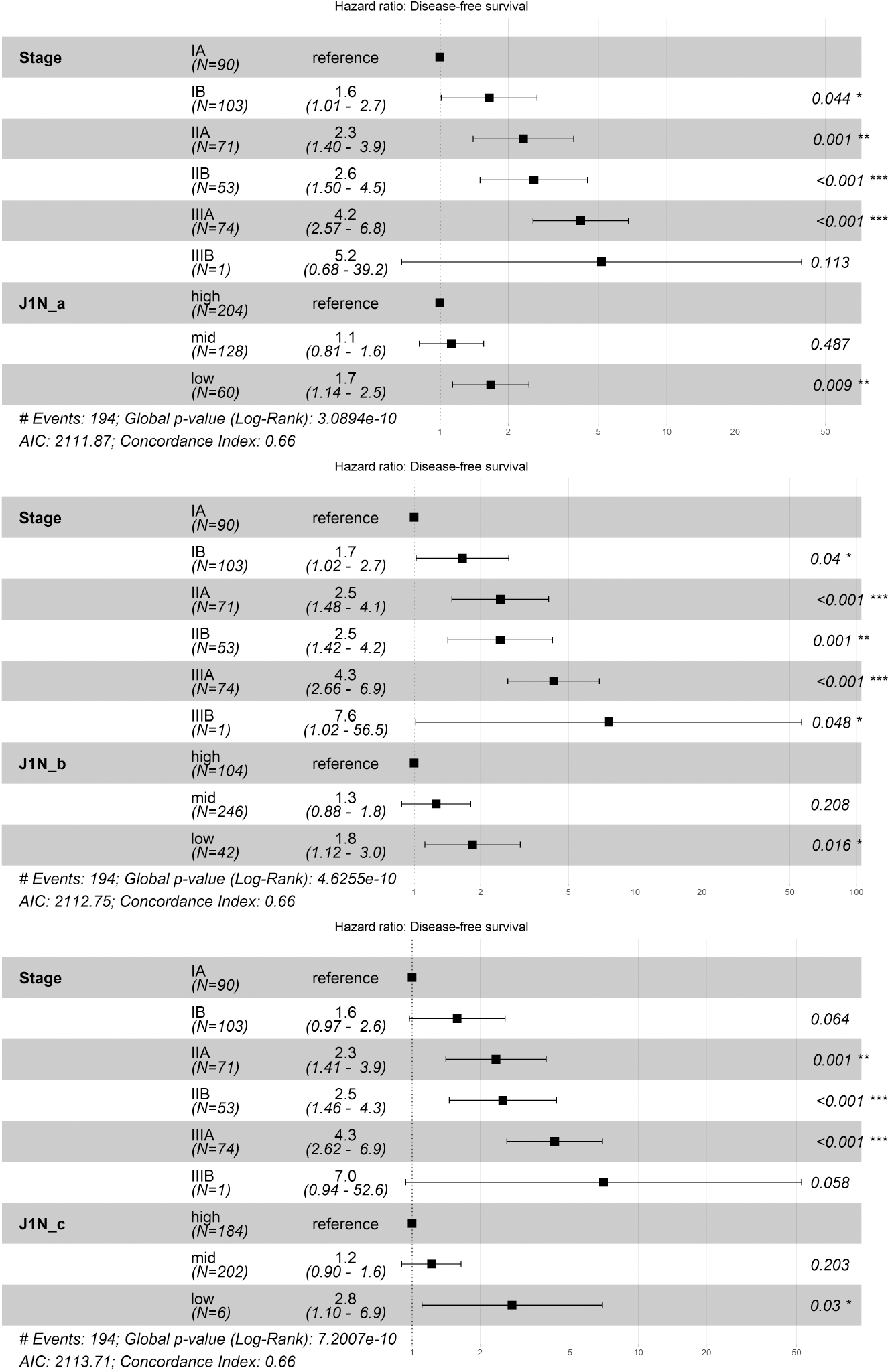
Multi-variable Cox proportional hazard models containing stage and alternative tree balance indices using the original cutpoints of 0.85 and 0.99. The HR 95% CIs are shown in brackets and by the error bars. The asterisks indicate the *P* value ranges, where \**P* < 0.05, \*\**P* < 0.01, \*\*\**P* < 0.001.

**Figure 15:**
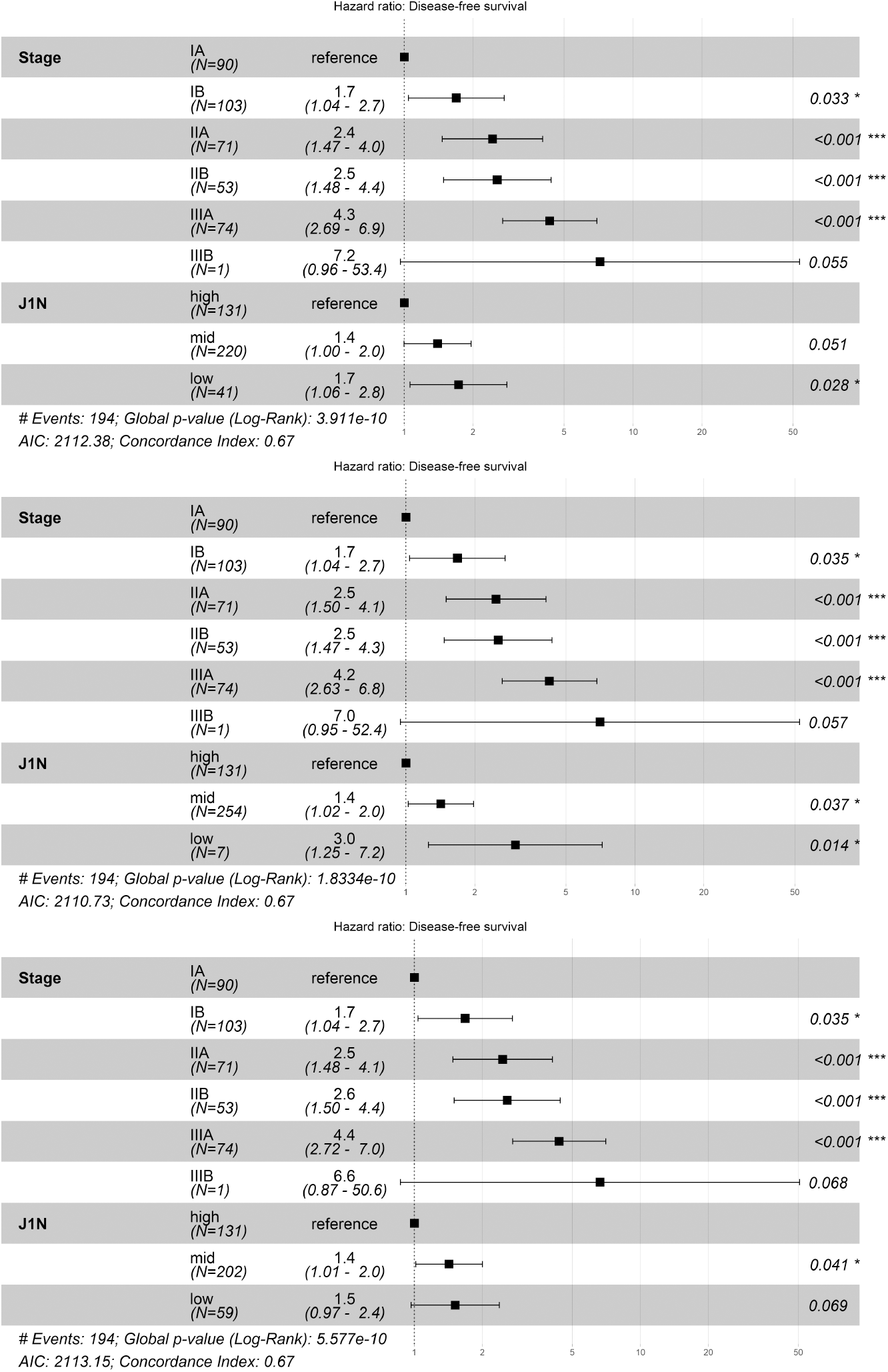
Multi-variable Cox proportional hazard models containing stage and the tree balance index, ^1^*J*_*N*_. The lower cutpoints here are chosen such that they give a “low” size group as close to the groupings for the alternative indices with the original cut points (Figure 14. The HR 95% CIs are shown in brackets and by the error bars. The asterisks indicate the *P* value ranges, where \**P* < 0.05, \*\**P* < 0.01, \*\*\**P* < 0.001.

## Notes

### Competing Interest Statement

The authors have declared no competing interest.

### Funding Statement

This study did not receive any funding

### Author Declarations

The study used ONLY openly available human data that were originally located at https://zenodo.org/records/7822002. This data set is associated with Frankell et al. (2023). The evolution of lung cancer and impact of subclonal selection in TRACERx. Nature, 616(7957), 525-533.

### Summary of Updates

Cited a few more prior studies in the Introduction. Improved Figure 2 to show trees with proportional branch lengths and node sizes. Added Results subsection "Associations between tree balance and clinical features". Added comparison with Shannon diversity (Figure 8f).

